# Time-varying and tissue-dependent effects of adiposity on leptin levels: a Mendelian randomization study

**DOI:** 10.1101/2022.11.29.22282906

**Authors:** Tom G Richardson, Genevieve M Leyden, George Davey Smith

## Abstract

**Background:** Findings from Mendelian randomization (MR) studies are conventionally interpreted as lifelong effects, which typically do not provide insight into the molecular mechanisms underlying the effect of an exposure on an outcome. In this study, we apply two recently developed MR approaches (known as ‘lifecourse’ and ‘tissue-partitioned’ MR) to investigate lifestage specific effects and tissues of action in the relationship between adiposity and circulating leptin levels.

**Method:** Genetic instruments for childhood and adult adiposity were used to estimate lifestage specific effects on leptin levels measured during early life (mean age:10 years) in the Avon Longitudinal Study of Parents and Children (ALSPAC) and in adulthood (mean age:55 years) using summary-level data from the deCODE Health study. This was followed by partitioning body mass index (BMI) instruments into those whose effects are putatively mediated by gene expression in either subcutaneous adipose or brain tissues, followed by using MR to simultaneously estimate their separate effects on childhood and adult leptin levels.

**Results:** There was strong evidence that childhood adiposity has a direct effect on leptin levels at age 10 years in the lifecourse (Beta=1.10, 95% CI=0.90 to 1.30, P=6×10^−28^), whereas evidence of an indirect effect was found on adulthood leptin along the causal pathway involving adulthood body size (Beta=0.74, 95% CI=0.62 to 0.86, P=1×10^−33^). Tissue-partitioned MR analyses provided evidence to suggest that BMI exerts its effect on leptin levels during both childhood and adulthood via brain-tissue mediated pathways (Beta=0.79, 95% CI=0.22 to 1.36, P=6×10^−3^ & Beta=0.51, 95% CI=0.32 to 0.69, P=1×10^−7^ respectively).

**Conclusions:** Our findings demonstrate the use of lifecourse MR to disentangle direct and indirect effects of early life exposures on time-varying complex outcomes. Furthermore, by integrating tissue-specific data, we highlight the aetiological importance of appetite regulation in the effect of adiposity on leptin levels, as well as raising implications for the gene-environment equivalence assumption in MR.

**Funding:** This work was supported by the Integrative Epidemiology Unit which receives funding from the UK Medical Research Council and the University of Bristol (MC_UU_00011/1).

## Introduction

People living with obesity have elevated levels of the peptide hormone leptin. This can be attributed to the amount of leptin in circulation being proportional to the amount of adipose tissue that an individual has [1]. After being secreted by fat cells in adipose tissue, leptin predominantly acts in the hypothalamus as a major regulator of energy balance [2]. Likewise, both neural and adipose tissues are known to play an important role in the molecular aetiology of body mass index (BMI), which is conventionally used to clinically diagnose obesity. However, despite being a cost-effective approach to routinely measure adiposity at scale, BMI is a construct which captures multiple heterogeneous subcomponents. This is epitomized by previous investigations into the functional genes which exert their effects on adiposity via their expression in brain and adipose tissues [3, 4], highlighting the divergent pathways that exist between BMI and downstream complex traits.

Mendelian randomization (MR) is a causal inference technique that can exploit the random segregation of genetic variants within a population to evaluate the genetically predicted effects of modifiable exposures on complex outcomes [5, 6]. We recently extended the principles of MR to evaluate the separate effects of molecular subcomponents of BMI using genetic variants partitioned by their impact on gene expression derived from subcutaneous adipose and brain tissue (known as ‘tissue-partitioned MR’) [7]. Using this approach suggested that the brain-derived variants are predominantly responsible for driving the effect of BMI on cardiometabolic disease outcomes [7], which we postulate is due to their involvement in appetite regulation and energy expenditure. Conversely, the subcutaneous adipose-derived set of instruments were predominantly responsible for the effect of BMI on outcomes such as endometrial cancer [8].

In this study, we sought to dissect the causal pathway between BMI and circulating leptin using tissue-partitioned MR by leveraging these brain- and adipose-tissue derived sets of instruments. However, investigating this hypothesis is made even more challenging given that associations between BMI and leptin levels have been reported as early in life as childhood [9]. Therefore, to further develop insight into the relationship between BMI and circulating leptin, we additionally applied another extension which we have developed in recent years known as ‘lifecourse MR’ [10]. This approach allows the independent effects of childhood and adult adiposity to be simultaneously estimated on an outcome which can also be measured at separate timepoints in the lifecourse [11, 12], such as childhood (mean age: 10 years) and adulthood (mean age: 55 years) measures of circulating leptin levels in this study. Taken together, we aimed to conduct a proof-of-concept study for these novel extensions of the conventional MR approach.

## Materials and Methods

### Childhood and adulthood measures of circulating leptin

The Avon Longitudinal Study of Parents and Children (ALSPAC) is a population-based cohort investigating genetic and environmental factors that affect the health and development of children. The study methods are described in detail elsewhere [13, 14]. In brief, 14,541 pregnant women residents in the former region of Avon, UK, with an expected delivery date between April 1, 1991 and December 31, 1992, were eligible to take part in ALSPAC. Of these initial pregnancies, there was a total of 14,676 foetuses, resulting in 14,062 live births and 13,988 children who were alive at 1 year of age. Please note that the study website contains details of all the data that is available through a fully searchable data dictionary and variable search tool” and reference the following webpage: http://www.bristol.ac.uk/alspac/researchers/our-data/. Written informed consent was obtained for all study participants. Ethical approval for this study was obtained from the ALSPAC Ethics and Law Committee and the Local Research Ethics Committees. Childhood measures of circulating leptin levels were obtained from non-fasting blood samples taken from ALSPAC participants at mean age 9.9 years (range=8.9 to 11.5 years). Analyses in ALSPAC were undertaken on a final sample size of 4,155 individuals after removing those without genetic data and withdrawn consent.

### Adulthood estimates of circulating leptin levels and fat distribution

Genetic estimates on adulthood circulating leptin levels were obtained from a previously conducted large-scale study of 35,559 individuals enrolled in the deCODE Health study (mean age: 55 years, standard deviation: 17 years). Full details on that study have been reported previously [15]. Briefly, circulating leptin was measured from plasma samples using the SomaScan version 4 assay by SomaLogic. Previously conducted genetic analyses identified a strong association between carriers of a missense variant (rs774055408) located in the leptin gene (*LEP*) and this measure of circulating leptin (P=6.2×10^−12^), reinforcing its validity for downstream analyses such as those undertaken in this study. This assay has also been reported to provide a very highly correlated measure of circulating leptin using the antibody based Olink platform (r=0.95) [16]. All participants from this study who donated samples gave informed consent, and the National Bioethics Committee of Iceland approved the study, which was conducted in agreement with conditions issued by the Data Protection Authority of Iceland (VSN_14-015). Genome-wide effect estimates on measures of fat distribution were obtained from a recent study on 38,965 UK Biobank participants who analysed MRI-derived measures of visceral, abdominal subcutaneous and gluteofemoral fat tissue volumes [17].

### Lifecourse Mendelian randomization

Lifecourse MR has been described in detail previously [10] and further details can be found in **Supplementary Note 1**. Briefly, genetic instruments derived in the UK Biobank study [10] have been shown to separate clinically measured childhood and adult BMI in 3 independent cohorts (ALSPAC [10], the Young Finns Study [18] and the Trøndelag Health (HUNT) study [19]). Both univariable and multivariable MR analyses on childhood leptin were conducted in a one-sample setting using individual level data from ALSPAC by analysing genetic risk scores with adjustment for age and sex. Univariable MR analyses to estimate total effects on adulthood leptin were undertaken in a two-sample setting using the inverse variance weighted (IVW) method [20], as well as the weighted median and MR-Egger methods [21, 22]. Multivariable MR analyses on adulthood leptin were also performed in a two-sample to estimate the direct and indirect effects of childhood body size [23].

### Tissue-partitioned Mendelian randomization

Instrument derivation and methodology for tissue partitioned MR has also been described in detail previously [7]. In brief, 86 independent BMI-associated instruments provided evidence of sharing a causal variant with proximal gene expression in subcutaneous adipose tissue through genetic colocalization analyses (based on a posterior probability (PPA)>0.8). The same approach provided evidence that the effects of 140 BMI instruments are putatively mediated by a nearby gene’s expression in brain tissue. These two sets of genetic variants have near identical average effect sizes on BMI (adipose=0.0148 and brain=0.0149 standard deviation change in BMI per effect allele), although the manner in which they relate to other anthropometric traits can markedly differ (e.g. waist-to-hip ratio, where adipose variants Pearson correlation coefficient (r)=0.45 & brain variants r=0.73. Univariable and multivariable MR analyses on childhood leptin measured in ALSPAC were undertaken as above in a one-sample setting by aggregating the tissue-partitioned BMI instruments as genetic risk scores and analysing childhood leptin with adjustment for age and sex. Estimates on adulthood leptin were initially evaluated in a two-sample setting using the three univariable MR methods, and subsequently using an extension of multivariable MR with instruments weighted by their PPA values for each tissue type [8]. Further details on this approach can be found in **Supplementary Note 2**. An overview of datasets used for exposures and outcomes analysed in this study can be found in **Supplementary Table 1**. All analyses were conducted using the ‘TwoSampleMR’ R package [24].

## Results

### Disentangling direct and indirect effects of childhood adiposity on early and mid-life measures of leptin levels

We firstly estimated the total effect of childhood adiposity on circulating leptin using data measured in ALSPAC participants at mean age 9.9 years in the lifecourse (Beta=1.28 per 1-standard deviation (SD) change in leptin levels per change in body size category, 95% CI=1.10 to 1.46, P=2×10^−41^). This was followed by applying multivariable MR which provided evidence that childhood body size has a direct effect on increased leptin levels at this point in the lifecourse (Beta=1.10, 95% CI=0.90 to 1.30, P=6×10^−28^) (**Figure 1A & Figure 1B, Supplementary Table 2**). Both childhood and adult adiposity likewise provided evidence of a total effect on circulating leptin measured in adulthood (Beta=0,70, 95% CI=0.58 to 0.81, P=6×10^−33^ & Beta=0.87, 95% CI=0.79 to 0,95, P=6×10^−93^ respectively) (**Supplementary Table 3**). However, multivariable MR analyses suggested that childhood adiposity indirectly influences leptin levels measured during adulthood along the causal pathway involving adult body size (Beta=0.74, 95% CI=0.62 to 0.86, P=1×10^−33^) (**Figure 1C & Figure 1D, Supplementary Table 4**).

**Figure 1.**
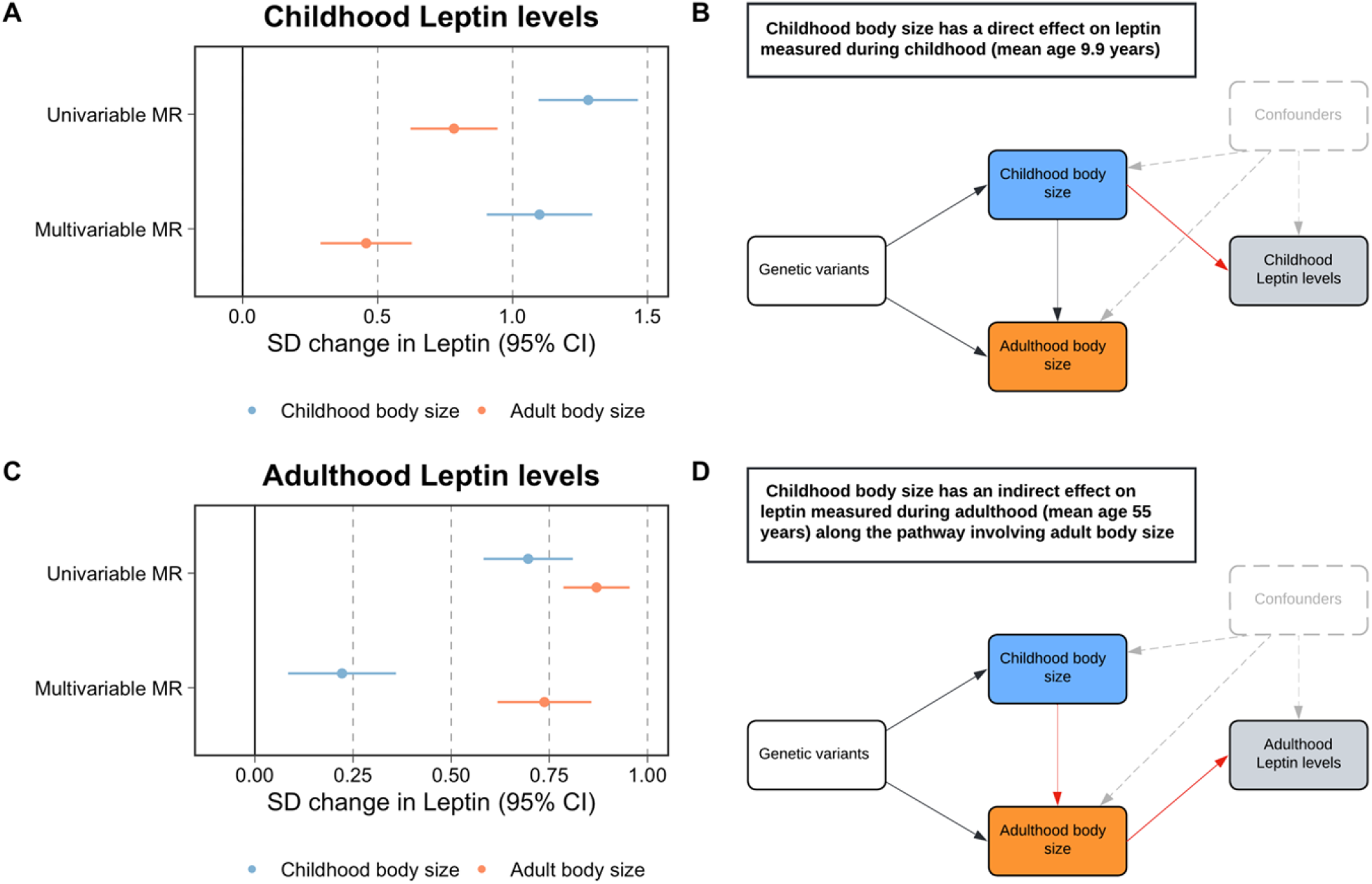
A) A forest plot which illustrates the direct effect of childhood body size on circulating leptin levels measured during childhood (mean age: 9.9 years) using individual-level data from the Avon Longitudinal Study of Parents and Children (ALSPAC) B) provides the corresponding schematic diagram for ALSPAC results C) A forest plot depicting the indirect effect of childhood body size on adulthood measured leptin levels using data from the deCODE Health study (mean age: 55 years) as portrayed in the schematic diagram presented in panel D). MR – Mendelian randomization. Data underlying this figure can be found in Supplementary Tables 3 & 4.

### Separating the tissue-partitioned effects of body mass index on leptin levels measured during childhood and adulthood

Univariable MR analyses provided evidence of a total effect of adiposity on leptin levels measured during childhood based on analyses using the adipose-tissue (Beta=0.61, 95% CI=0.13 to 1.08, P=0.01) and brain-tissue (Beta=0.86, 95% CI=0.40 to 1.31, P=2×10^−4^) partitioned instruments. In a multivariable setting, the brain-tissue derived component of BMI predominated in the model (Beta=0.79, 95% CI=0.22 to 1.36, P=6×10^−3^), whereas the adipose-tissue derived estimate attenuated to include the null (Beta=0.12, 95% CI=-0.48 to 0.71, P=0.70) (**Figure 2A & Figure 2B, Supplementary Table 5**). Analyses on adulthood measured leptin also provided strong evidence of a total effect based on adipose-and brain-tissue derived estimates (Beta=0.52, 95% CI=0.39 to 0.65, P=2×10^−14^ & Beta=0.61, 95% CI=0.50 to 0.73, P=1×10^−25^ respectively). Similar to findings for childhood leptin, subcutaneous adipose-tissue derived estimates attenuated substantially more (Beta=0.23, 95% CI=0.01 to 0.45, P=0.04) compared to the estimates derived using the brain-tissue instrument set (Beta=0.51, 95% CI=0.32 to 0.69, P=1×10^−7^) in a multivariable setting (**Figure 2C & Figure 2D, Supplementary Tables 6 & 7**). As a further sensitivity analysis to characterize the causal pathway between these tissue-partitioned variants and fat distribution, we found that in particular the brain expressed instruments have a predominating effect on visceral fat volume (Beta=0.51, 95% CI=0.30 to 0.72, P=2×10^−6^), compared to the subcutaneous adipose instruments (Beta=0.07, 95% CI=-0.18 to 0.32, P=0.57) (**Supplementary Figure 1 & Supplementary Table 8**).

**Figure 2.**
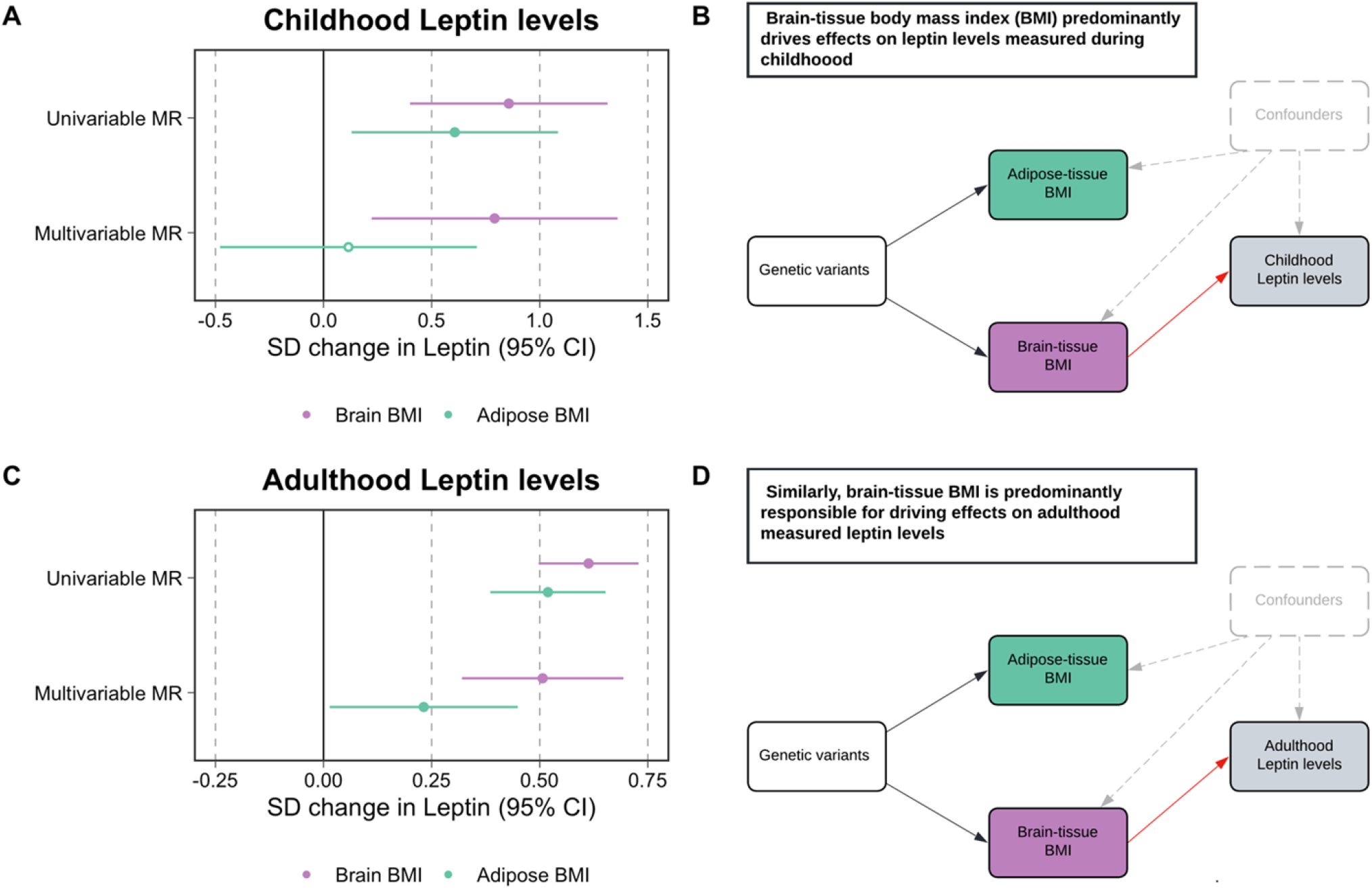
A) A forest plot illustrating the attenuation of the adipose-tissue instrumented body mass index (BMI) effect on childhood leptin levels, whereas estimates for brain-tissue instrumented BMI remained robust B) The corresponding schematic diagram for this finding C) A forest plot displaying similar conclusions to the estimates found in childhood, but derived using adulthood measured leptin levels. D) provides the corresponding schematic diagram for estimates portrayed in panel C). MR – Mendelian randomization. Data underlying this figure can be found in Supplementary Tables 6 & 7.

## Discussion

In this study, we applied two recent approaches in the MR paradigm, known as lifecourse and tissue partitioned MR, to investigate the influence of adiposity on circulating leptin levels as an exemplar relationship to demonstrate the value of these techniques. Whilst there is irrefutable evidence that having a higher BMI increases circulating levels of leptin, these two extensions of MR methodology provide insight into lifecourse-specific effects and the tissues of action underlying this aetiological relationship. We propose that these approaches can be conducted to build upon findings by conventional MR analyses, which use naturally occurring genetic variants as causal anchors to help establish and estimate the causal effect of modifiable exposure on complex outcomes and disease endpoints.

Our application of lifecourse MR in this work supports a breadth of previous research using this technique, which highlights the importance of taking into consideration the age at which data from participants for exposure and outcome datasets are analysed. Findings from this study provide evidence that adiposity in childhood exerts a direct effect on leptin levels when measured in early life, corroborating findings from observational studies [9]. Conversely, evidence of an indirect effect along the causal pathway involving adulthood adiposity was found when analysing leptin measured in later life. This provides a powerful proof of concept for this approach, which has previously found evidence of a direct effect of childhood adiposity on outcomes such as breast cancer [10], type 1 diabetes [25] and cardiac structure [26]. Evidence supporting indirect effects, as in this study for adulthood leptin, are likely attributed to individuals in a population remaining overweight for many years in the lifecourse, as found previously for various cardiovascular endpoints [27].

Tissue-partitioned MR analyses in this work suggest that the subcomponent of BMI proxied using genetic variants whose effects are putatively mediated via gene expression in brain tissue are predominantly responsible for driving effects of adiposity on leptin during both childhood and adulthood. We postulate that these findings highlight a role for appetite regulation and energy expenditure mechanisms as of fundamental importance in the effect of adiposity on leptin levels, putatively mediated along the causal pathway involving increased visceral fat storage. This corroborates findings from the literature, which suggest that being overweight has a downstream consequence on elevated leptin levels due to leptin resistance occurring in the central nervous system, particularly the hypothalamus [28]. This is influenced by factors such as blood-brain barrier permeability, which results in leptin failing to suppress appetite and consequently leads to circulating hyperleptinemia amongst patients with obesity [29].

Fractionating genetic instruments for the same exposure using molecular datasets has important considerations for the (often overlooked) gene-environment equivalence assumption in MR [30]. This states that the effect of germline genetic perturbations should have the same downstream consequence on outcomes as if they were caused by the modifiable exposures themselves. For the adipose- and brain-tissue partitioned sets of instruments used in this study, we found previously that the manner in which they relate to downstream disease and complex traits can drastically differ despite have almost identical average effect estimates on BMI as an exposure. In particular, this finding underlines the heterogeneous nature of BMI as a lifestyle risk factor and highlights that this human-derived construct likely captures various causal pathways underlying the relationship between anthropometry and complex outcomes. We emphasise that future application of tissue-partitioned MR should carefully consider both the exposure and functionally relevant tissue types being investigated, as well as ensuring that the derived instrument sets are equally predictive of the exposure [8].

We note that both lifecourse and tissue-partitioned MR have important caveats. For instance, the childhood and adult body size instruments used to disentangle direct and indirect effects in this study do not provide insight into other timepoints over the lifecourse (e.g. adolescence). Future efforts should focus on deriving instruments to separate effects of age-specific adiposity at more granular windows over the lifecourse. In addition, tissue-partitioned instruments were derived using bulk tissue in this work due to availability of data and therefore do not take into account cell-type heterogeneity [31, 32]. Furthermore, as with all estimates derived from MR, triangulating findings from other orthogonal lines of evidence derived using different study design and datasets provide the most robust conclusions for the approaches applied in this work [33].

In summary, our findings highlight a putative role for genes expressed in neural tissues in the aetiology of adiposity and leptin levels during both childhood and adulthood. Furthermore, this innovative study provides a proof-of-concept into how the principles of Mendelian randomization can be adapted to investigate hypotheses outside the scope of how this causal inference technique was originally conceived.

## Data Availability

All individual level data analysed in this study can be accessed via an approved application to ALSPAC (http://www.bristol.ac.uk/alspac/researchers/access/). Summary-level data on adulthood leptin levels were provided by the deCODE Health study which can be found at (https://www.decode.com/summarydata/).

## Data availability

All individual level data analysed in this study was obtained from the ALSPAC study which is not allowed to be deposited in a public repository. However, all data can be accessed via an application to ALSPAC which requires approval from executive committee (http://www.bristol.ac.uk/alspac/researchers/access/). Summary-level data on adulthood leptin levels were provided by the deCODE Health study which can be found at (https://download.decode.is/form/folder/proteomics). All other data analysed in this study is based on summary-level results as referenced throughout.

## Consent

Written informed consent was obtained for all study participants. Ethical approval for this study was obtained from the ALSPAC Ethics and Law Committee and the Local Research Ethics Committees.

## Competing Interests

TGR is employed by GlaxoSmithKline outside of this work. All other authors declare no conflicts of interest.

## Funding

This work was supported by the Integrative Epidemiology Unit which receives funding from the UK Medical Research Council and the University of Bristol (MC_UU_00011/1).

## Acknowledgements

We are extremely grateful to all the families who took part in this study, the midwives for their help in recruiting them and the whole ALSPAC team, which includes interviewers, computer and laboratory technicians, clerical workers, research scientists, volunteers, managers, receptionists and nurses. The UK Medical Research Council and Wellcome (Grant ref: 217065/Z/19/Z) and the University of Bristol provide core support for ALSPAC. Consent for biological samples has been collected in accordance with the Human Tissue Act (2004). GWAS data was generated by Sample Logistics and Genotyping Facilities at Wellcome Sanger Institute and LabCorp (Laboratory Corporation of America) using support from 23andMe.

This research was conducted at the NIHR Biomedical Research Centre at the University Hospitals Bristol NHS Foundation Trust and the University of Bristol. The views expressed in this publication are those of the author(s) and not necessarily those of the NHS, the National Institute for Health Research or the Department of Health. This publication is the work of the authors and TGR will serve as guarantor for the contents of this paper.

---

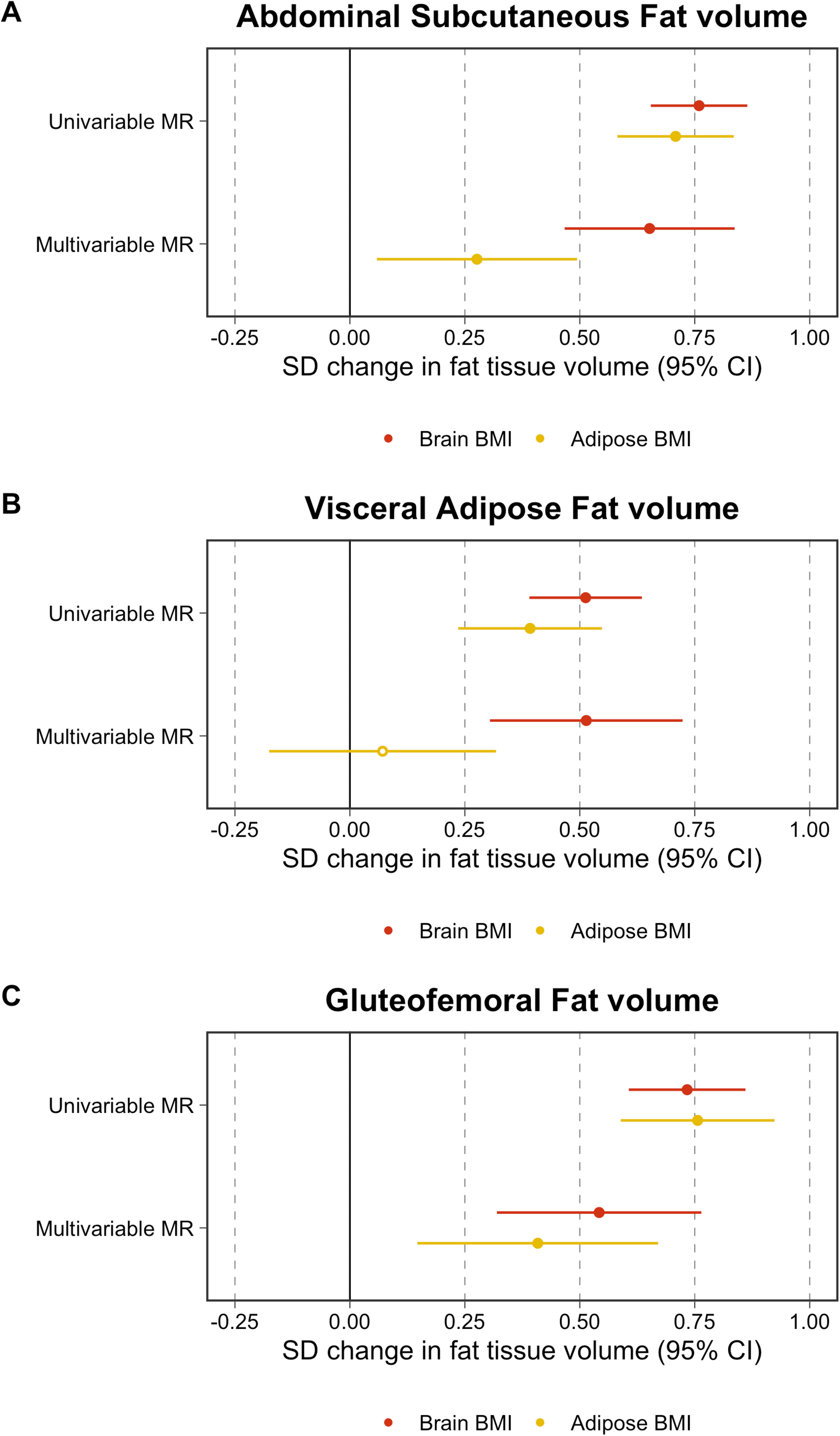

## Notes

### Competing Interest Statement

TGR is an employee of GlaxoSmithKline outside of this work.

## References

1. Obradovic M, Sudar-Milovanovic E, Soskic S, Essack M, Arya S, Stewart AJ, et al. Leptin and Obesity: Role and Clinical Implication. Front Endocrinol (Lausanne). 2021;12:585887. Epub 2021/06/05. doi: 10.3389/fendo.2021.585887. PubMed PMID: 34084149; PubMed Central PMCID: PMCPMC8167040.

2. Morrison CD. Leptin signaling in brain: A link between nutrition and cognition? Biochim Biophys Acta. 2009;1792(5):401–8. Epub 2009/01/10. doi: 10.1016/j.bbadis.2008.12.004. PubMed PMID: 19130879; PubMed Central PMCID: PMCPMC2670357.

3. Timshel PN, Thompson JJ, Pers TH. Genetic mapping of etiologic brain cell types for obesity. Elife. 2020;9. Epub 2020/09/22. doi: 10.7554/eLife.55851. PubMed PMID: 32955435; PubMed Central PMCID: PMCPMC7505664.

4. Rask-Andersen M, Karlsson T, Ek WE, Johansson A. Genome-wide association study of body fat distribution identifies adiposity loci and sex-specific genetic effects. Nat Commun. 2019;10(1):339. Epub 2019/01/22. doi: 10.1038/s41467-018-08000-4. PubMed PMID: 30664634; PubMed Central PMCID: PMCPMC6341104.

5. Davey Smith G, Ebrahim S. ‘Mendelian randomization’: can genetic epidemiology contribute to understanding environmental determinants of disease? Int J Epidemiol. 2003;32(1):1–22. Epub 2003/04/12. doi: 10.1093/ije/dyg070. PubMed PMID: 12689998.

6. Richmond RC, Davey Smith G. Mendelian Randomization: Concepts and Scope. Cold Spring Harb Perspect Med. 2022;12(1). Epub 2021/08/25. doi: 10.1101/cshperspect.a040501. PubMed PMID: 34426474; PubMed Central PMCID: PMCPMC8725623.

7. Leyden GM, Shapland CY, Davey Smith G, Sanderson E, Greenwood MP, Murphy D, et al. Harnessing tissue-specific genetic variation to dissect putative causal pathways between body mass index and cardiometabolic phenotypes. Am J Hum Genet. 2022;109(2):240–52. Epub 2022/01/30. doi: 10.1016/j.ajhg.2021.12.013. PubMed PMID: 35090585; PubMed Central PMCID: PMCPMC8874216.

8. Leyden GM, Greenwood MP, Gaborieau V, Younghun H, Amos C, Brennan P, et al. Disentangling the aetiological pathways between body mass index and site-specific cancer risk using tissue-partitioned Mendelian randomization. Under review. 2022.

9. Shalitin S, Phillip M. Role of obesity and leptin in the pubertal process and pubertal growth--a review. Int J Obes Relat Metab Disord. 2003;27(8):869–74. Epub 2003/07/16. doi: 10.1038/sj.ijo.0802328. PubMed PMID: 12861226.

10. Richardson TG, Sanderson E, Elsworth B, Tilling K, Davey Smith G. Use of genetic variation to separate the effects of early and later life adiposity on disease risk: mendelian randomisation study. BMJ. 2020;369:m1203. Epub 2020/05/08. doi: 10.1136/bmj.m1203. PubMed PMID: 32376654; PubMed Central PMCID: PMCPMC7201936 at www.icmje.org/coi_disclosure.pdf and declare: funding by grants from the Medical Research Council and Health Data Research UK for the submitted work; no support from any other organisation for the submitted work; no financial relationships with any other organisations that might have an interest in the submitted work in the previous three years; no other relationships or activities that could appear to have influenced the submitted work.

11. Richardson TG, Power GM, Davey Smith G. Adiposity may confound the association between vitamin D and disease risk - a lifecourse Mendelian randomization study. Elife. 2022;11. Epub 2022/08/09. doi: 10.7554/eLife.79798. PubMed PMID: 35938910; PubMed Central PMCID: PMCPMC9359699.

12. Sanderson E, Richardson TG, Morris TT, Tilling K, Smith GD. Estimation of causal effects of a time-varying exposure at multiple time points through multivariable mendelian randomization. PLoS Genet. 2022;18(7):e1010290. Epub 2022/07/19. doi: 10.1371/journal.pgen.1010290. PubMed PMID: 35849575.

13. Boyd A, Golding J, Macleod J, Lawlor DA, Fraser A, Henderson J, et al. Cohort Profile: the ‘children of the 90s’--the index offspring of the Avon Longitudinal Study of Parents and Children. Int J Epidemiol. 2013;42(1):111–27. Epub 2012/04/18. doi: 10.1093/ije/dys064. PubMed PMID: 22507743; PubMed Central PMCID: PMCPMC3600618.

14. Fraser A, Macdonald-Wallis C, Tilling K, Boyd A, Golding J, Davey Smith G, et al. Cohort Profile: the Avon Longitudinal Study of Parents and Children: ALSPAC mothers cohort. Int J Epidemiol. 2013;42(1):97–110. Epub 2012/04/18. doi: 10.1093/ije/dys066. PubMed PMID: 22507742; PubMed Central PMCID: PMCPMC3600619.

15. Ferkingstad E, Sulem P, Atlason BA, Sveinbjornsson G, Magnusson MI, Styrmisdottir EL, et al. Large-scale integration of the plasma proteome with genetics and disease. Nat Genet. 2021;53(12):1712–21. Epub 2021/12/04. doi: 10.1038/s41588-021-00978-w. PubMed PMID: 34857953.

16. Pietzner M, Wheeler E, Carrasco-Zanini J, Kerrison ND, Oerton E, Koprulu M, et al. Synergistic insights into human health from aptamer- and antibody-based proteomic profiling. Nat Commun. 2021;12(1):6822. Epub 2021/11/26. doi: 10.1038/s41467-021-27164-0. PubMed PMID: 34819519; PubMed Central PMCID: PMCPMC8613205.

17. Agrawal S, Wang M, Klarqvist MDR, Smith K, Shin J, Dashti H, et al. Inherited basis of visceral, abdominal subcutaneous and gluteofemoral fat depots. Nat Commun. 2022;13(1):3771. Epub 2022/07/01. doi: 10.1038/s41467-022-30931-2. PubMed PMID: 35773277; PubMed Central PMCID: PMCPMC9247093.

18. Richardson TG, Mykkanen J, Pahkala K, Ala-Korpela M, Bell JA, Taylor K, et al. Evaluating the direct effects of childhood adiposity on adult systemic metabolism: a multivariable Mendelian randomization analysis. Int J Epidemiol. 2021. Epub 2021/03/31. doi: 10.1093/ije/dyab051. PubMed PMID: 33783488.

19. Brandkvist M, Bjorngaard JH, Odegard RA, Asvold BO, Smith GD, Brumpton B, et al. Separating the genetics of childhood and adult obesity: a validation study of genetic scores for body mass index in adolescence and adulthood in the HUNT Study. Hum Mol Genet. 2020. Epub 2020/12/05. doi: 10.1093/hmg/ddaa256. PubMed PMID: 33276378.

20. Burgess S, Butterworth A, Thompson SG. Mendelian randomization analysis with multiple genetic variants using summarized data. Genet Epidemiol. 2013;37(7):658–65. Epub 2013/10/12. doi: 10.1002/gepi.21758. PubMed PMID: 24114802; PubMed Central PMCID: PMCPMC4377079.

21. Bowden J, Davey Smith G, Burgess S. Mendelian randomization with invalid instruments: effect estimation and bias detection through Egger regression. Int J Epidemiol. 2015;44(2):512–25. Epub 2015/06/08. doi: 10.1093/ije/dyv080. PubMed PMID: 26050253; PubMed Central PMCID: PMCPMC4469799.

22. Bowden J, Davey Smith G, Haycock PC, Burgess S. Consistent Estimation in Mendelian Randomization with Some Invalid Instruments Using a Weighted Median Estimator. Genet Epidemiol. 2016;40(4):304–14. Epub 2016/04/12. doi: 10.1002/gepi.21965. PubMed PMID: 27061298; PubMed Central PMCID: PMCPMC4849733.

23. Sanderson E, Davey Smith G, Windmeijer F, Bowden J. An examination of multivariable Mendelian randomization in the single-sample and two-sample summary data settings. Int J Epidemiol. 2019;48(3):713–27. Epub 2018/12/12. doi: 10.1093/ije/dyy262. PubMed PMID: 30535378.

24. Hemani G, Zheng J, Elsworth B, Wade KH, Haberland V, Baird D, et al. The MR-Base platform supports systematic causal inference across the human phenome. Elife. 2018;7. Epub 2018/05/31. doi: 10.7554/eLife.34408. PubMed PMID: 29846171; PubMed Central PMCID: PMCPMC5976434.

25. Richardson TG, Crouch DJM, Power GM, Morales-Berstein F, Hazelwood E, Fang S, et al. Childhood body size directly increases type 1 diabetes risk based on a lifecourse Mendelian randomization approach. Nat Commun. 2022;13(1):2337. Epub 2022/04/29. doi: 10.1038/s41467-022-29932-y. PubMed PMID: 35484151; PubMed Central PMCID: PMCPMC9051135.

26. O’Nunain K, Park C, Urquijo H, Leyden GM, Hughes AD, Davey Smith G, et al. A lifecourse mendelian randomization study highlights the long-term influence of childhood body size on later life heart structure. PLoS Biol. 2022;20(6):e3001656. Epub 2022/06/10. doi: 10.1371/journal.pbio.3001656. PubMed PMID: 35679339.

27. Power GM, Tyrrell J, Frayling TM, Davey Smith G, Richardson TG. Mendelian Randomization Analyses Suggest Childhood Body Size Indirectly Influences End Points From Across the Cardiovascular Disease Spectrum Through Adult Body Size. J Am Heart Assoc. 2021;10(17):e021503. Epub 2021/09/02. doi: 10.1161/JAHA.121.021503. PubMed PMID: 34465205; PubMed Central PMCID: PMCPMC8649247.

28. Gruzdeva O, Borodkina D, Uchasova E, Dyleva Y, Barbarash O. Leptin resistance: underlying mechanisms and diagnosis. Diabetes Metab Syndr Obes. 2019;12:191–8. Epub 2019/02/19. doi: 10.2147/DMSO.S182406. PubMed PMID: 30774404; PubMed Central PMCID: PMCPMC6354688.

29. Izquierdo AG, Crujeiras AB, Casanueva FF, Carreira MC. Leptin, Obesity, and Leptin Resistance: Where Are We 25 Years Later? Nutrients. 2019;11(11). Epub 2019/11/14. doi: 10.3390/nu11112704. PubMed PMID: 31717265; PubMed Central PMCID: PMCPMC6893721.

30. Davey Smith G. Epigenesis for epidemiologists: does evo-devo have implications for population health research and practice? Int J Epidemiol. 2012;41(1):236–47. Epub 2012/03/17. doi: 10.1093/ije/dys016. PubMed PMID: 22422459.

31. Glastonbury CA, Couto Alves A, El-Sayed Moustafa JS, Small KS. Cell-Type Heterogeneity in Adipose Tissue Is Associated with Complex Traits and Reveals Disease-Relevant Cell-Specific eQTLs. Am J Hum Genet. 2019;104(6):1013–24. Epub 2019/05/28. doi: 10.1016/j.ajhg.2019.03.025. PubMed PMID: 31130283; PubMed Central PMCID: PMCPMC6556877.

32. Prince C, Mitchell RE, Richardson TG. Integrative multiomics analysis highlights immune-cell regulatory mechanisms and shared genetic architecture for 14 immune-associated diseases and cancer outcomes. Am J Hum Genet. 2021;108(12):2259–70. Epub 2021/11/07. doi: 10.1016/j.ajhg.2021.10.003. PubMed PMID: 34741802; PubMed Central PMCID: PMCPMC8715275.

33. Munafo MR, Davey Smith G. Robust research needs many lines of evidence. Nature. 2018;553(7689):399–401. Epub 2018/01/26. doi: 10.1038/d41586-018-01023-3. PubMed PMID: 29368721.

